# Persistence of G10P[11] neonatal rotavirus infections in southern India

**DOI:** 10.1101/2021.03.01.21251320

**Authors:** Sudhir Babji, Kulandaipalayam Natarajan Sindhu, Sribal Selvarajan, Sasirekha Ramani, Srinivasan Venugopal, Shainey Alokit Khakha, Priya Hemavathy, Santhosh Kumar Ganesan, Sidhartha Giri, Sudhabharathi Reju, Krithika Gopalakrishnan, Binu Ninan, Miren Iturriza-Gomara, Padma Srikanth, Gagandeep Kang

**Affiliations:** The Wellcome Trust Research Laboratory, Division of Gastrointestinal Sciences, Christian Medical College, Vellore, India; Department of Microbiology, Sri Ramachandra Medical College and Research Institute, Chennai, India; Baylor College of Medicine, Houston, Texas, USA; Department of Neonatology, Sri Ramachandra Medical College and Research Institute, Chennai, India; NIHR Health Protection Research Unit in Gastrointestinal Infections at University of Liverpool, Liverpool, UK

**Keywords:** G10P[11] strain, neonate, nursery, rotavirus infection

## Abstract

Neonatal rotavirus infections are predominantly caused by distinct genotypes restricted to this age-group and are mostly asymptomatic. Stool samples from neonates admitted for >48 hours in neonatal intensive care units (NICUs) in Vellore (2014-2015) and Chennai (2015-2016) in southern India, and from neonates born at hospitals in Vellore but not admitted to NICUs (2015-2016) were tested for rotavirus by ELISA and genotyped by hemi-nested RT-PCR. Of the 791 neonates, 150 and 336 were recruited from Vellore and Chennai NICUs, and 305 were born in five hospitals in Vellore. The positivity rates in the three settings were 49.3% (74/150), 29.5% (99/336) and 54% (164/305), respectively. G10P[11] was the commonly identified genotype in 87.8% (65/74), 94.9% (94/99) and 98.2% (161/164) of the neonates in Vellore and Chennai NICUs, and those born at Vellore hospitals, respectively. Neonates delivered by lower segment caesarean section (LSCS) at Vellore hospitals, not admitted to NICUs, had a significantly higher odds of acquiring rotavirus infection compared to those delivered vaginally [*p=0*.*002*, OR=2.4 (1.4-4.3)]. This report demonstrates the persistence of G10P[11] strain in Vellore and Chennai, indicating widespread neonatal G10P[11] strain in southern India and their persistence over two decades, leading to interesting questions about strain stability.

---

Rotavirus infections in early infancy have been reported from both the developing and developed world (1). These infections in young infants can be both symptomatic and asymptomatic, with a large proportion of infants experiencing rotavirus infections at a much earlier age in low-income settings (2).Previous reports from Vellore, south India, described 56% of infants experiencing a rotavirus infection before they reach six months of age (3). Neonatal rotavirus infections have been reported from neonatal nurseries and intensive care units, as well as from the community (4–6).

There are 36 G and 51 P types of group A rotaviruses which have been associated with human infections (7). The most common genotypes associated with moderate to severe acute rotavirus infection in infants are G1P[8], G2P[4], G3P[8], G4P[8], G9P[8] and G12P[8], although temporal and spatial genotype distributions vary (8). Rotavirus genotypes specifically associated with neonatal infection are highly restricted to this age group and are rarely seen in older infants (9). Different genotypes have been reported in neonatal infections around the world (Table 1). Neonatal rotavirus infections are predominantly asymptomatic, although their association with gastrointestinal symptoms including necrotizing enterocolitis has been reported (10).

**TABLE 1.**
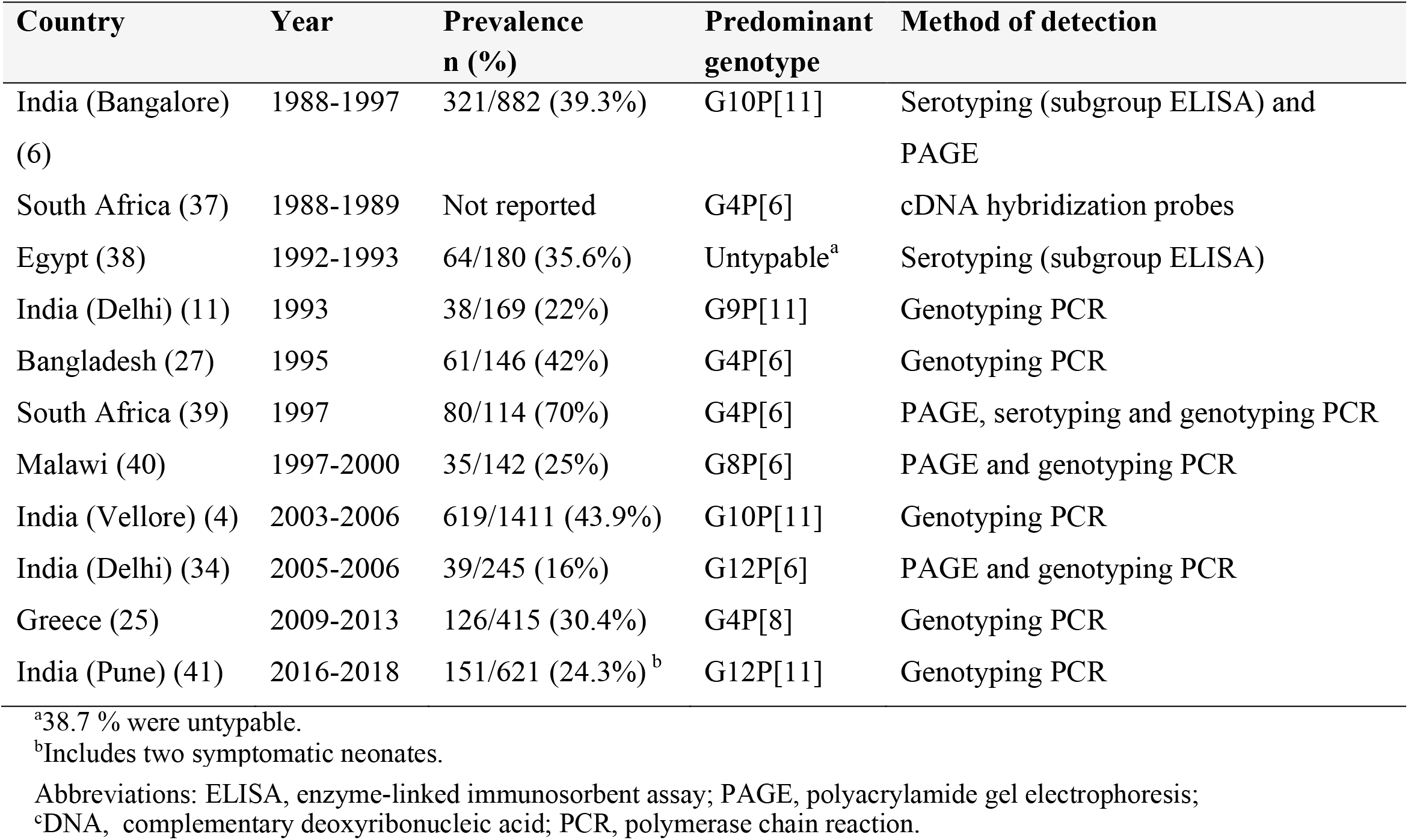
Neonatal rotavirus infections and strains reported from various countries between 1980 to 2096

Previously published reports from Vellore, south India, showed the predominance of the G10P[11] genotype in contrast to the G9P[11] genotype reported from Delhi, north India, or the G3P[6] from Australia (4,11,12). Prior studies in neonates from Vellore showed human milk oligosaccharides (HMO) increased the infectivity of the G10P[11] strain, by conferring structural stability to the strain, compared to the G1P[8] and G2[P4] genotypes, and potentially driving the persistence of the G10P[11] strains (13).

This study reports the prevalence of rotavirus infection and the strain type distribution in neonates in hospitals in Vellore and Chennai, pooling data from three studies.

## MATERIAL AND METHODS

Neonates from three studies were included. The first study enrolled neonates admitted for more than 48 hours to the neonatal intensive care unit (NICU) at Christian Medical College (CMC), Vellore during 2014-2015, for a study on the impact of HMO, breast milk microbiome and infant gut microbiome on neonatal rotavirus infections (13). Symptomatic and asymptomatic term and pre-term neonates were included in this study (Table 2 and 3). The second study included neonates enrolled from the NICU of Sri Ramachandra Medical College (SRMC), a tertiary care hospital in Chennai city, the state capital of Tamil Nadu, located ∼130 kilometres from Vellore. This study was conducted between 2015-2016. The third study included new-borns recruited in the Rotavirus Vaccine Immunogenicity study cohort (RoVI) between 2015-2016 from hospitals in and around Vellore namely, CMC, Vellore, the Low Cost-Effective Care Unit (LCECU), the Community Health and Development hospital (CHAD); the Government Vellore Medical College and hospital (GVMCH), and three Urban Health Centres (UHC) (14). A small number of infants were born at private nursing homes/hospitals. Only healthy infants not requiring hospitalisation for longer than 48 hours were included in the RoVI study cohort. Infants, born to mothers by LSCS had to stay in the hospital for longer with their mothers compared to those born vaginally. Written informed consent was obtained from the parent/legal guardian of the neonates from all the above cohorts.

**TABLE 2.**
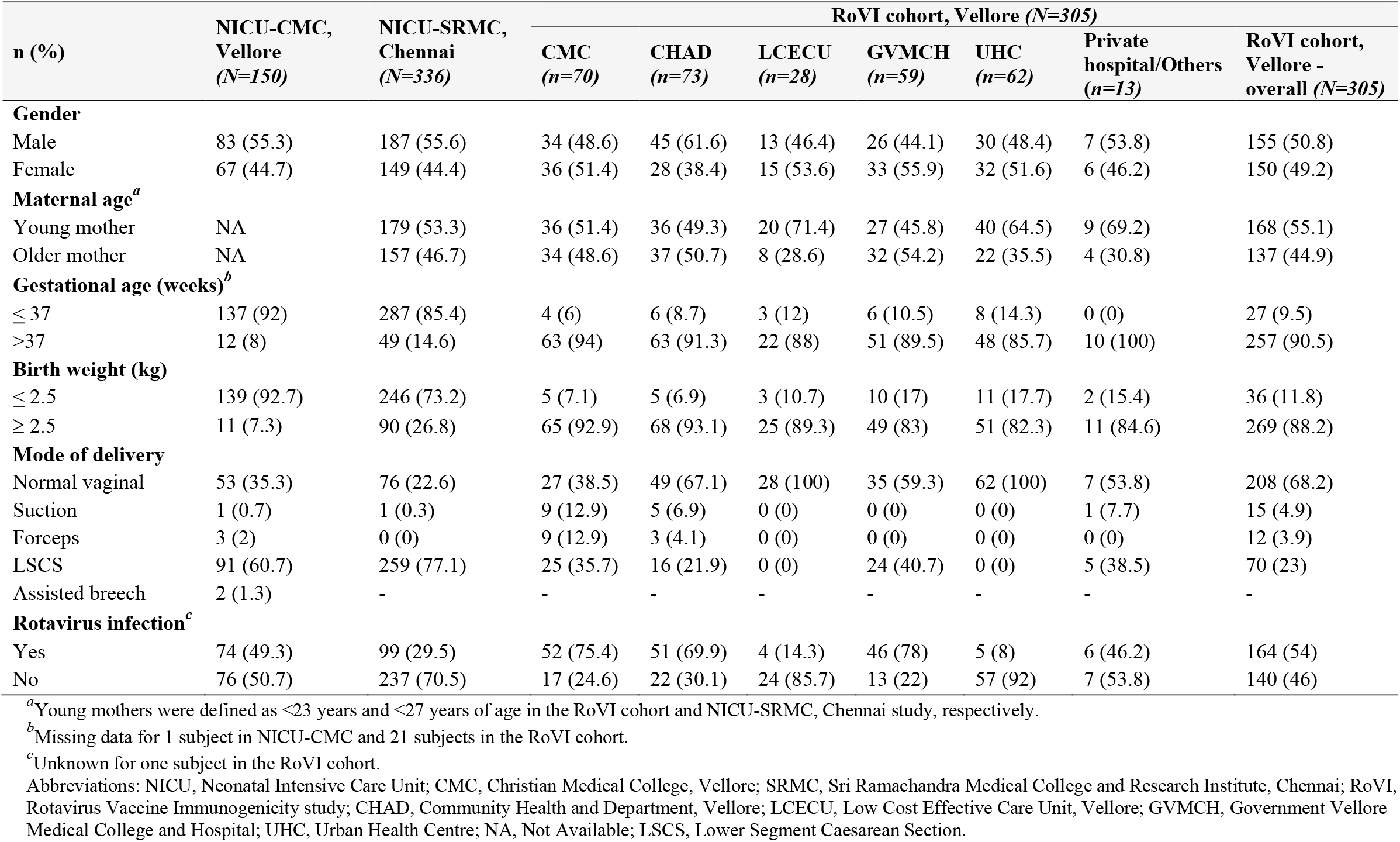
Baseline characteristics of the mother-infant pairs of the neonates from NICU-CMC, Vellore; NICU-SRMC and RoVI cohort

| n (%) | NICU-CMC,<br>Vellore<br>(N=150) | NICU-SRMC,<br>Chennai<br>(N=336) | RoVI cohort, Vellore (N=305) |  |  |  |  |  |  |
| --- | --- | --- | --- | --- | --- | --- | --- | --- | --- |
|  |  |  | CMC<br>(n=70) | CHAD<br>(n=73) | LCECU<br>(n=28) | GVMCH<br>(n=59) | UHC<br>(n=62) | Private<br>hospital/Others<br>(n=13) | RoVI cohort,<br>Vellore -<br>overall (N=305) |
| Gender |  |  |  |  |  |  |  |  |  |
| Male | 83 (55.3) | 187 (55.6) | 34 (48.6) | 45 (61.6) | 13 (46.4) | 26 (44.1) | 30 (48.4) | 7 (53.8) | 155 (50.8) |
| Female | 67 (44.7) | 149 (44.4) | 36 (51.4) | 28 (38.4) | 15 (53.6) | 33 (55.9) | 32 (51.6) | 6 (46.2) | 150 (49.2) |
| Maternal age<sup>a</sup> |  |  |  |  |  |  |  |  |  |
| Young mother | NA | 179 (53.3) | 36 (51.4) | 36 (49.3) | 20 (71.4) | 27 (45.8) | 40 (64.5) | 9 (69.2) | 168 (55.1) |
| Older mother | NA | 157 (46.7) | 34 (48.6) | 37 (50.7) | 8 (28.6) | 32 (54.2) | 22 (35.5) | 4 (30.8) | 137 (44.9) |
| Gestational age (weeks)<sup>b</sup> |  |  |  |  |  |  |  |  |  |
| ≤ 37 | 137 (92) | 287 (85.4) | 4 (6) | 6 (8.7) | 3 (12) | 6 (10.5) | 8 (14.3) | 0 (0) | 27 (9.5) |
| >37 | 12 (8) | 49 (14.6) | 63 (94) | 63 (91.3) | 22 (88) | 51 (89.5) | 48 (85.7) | 10 (100) | 257 (90.5) |
| Birth weight (kg) |  |  |  |  |  |  |  |  |  |
| ≤ 2.5 | 139 (92.7) | 246 (73.2) | 5 (7.1) | 5 (6.9) | 3 (10.7) | 10 (17) | 11 (17.7) | 2 (15.4) | 36 (11.8) |
| ≥ 2.5 | 11 (7.3) | 90 (26.8) | 65 (92.9) | 68 (93.1) | 25 (89.3) | 49 (83) | 51 (82.3) | 11 (84.6) | 269 (88.2) |
| Mode of delivery |  |  |  |  |  |  |  |  |  |
| Normal vaginal | 53 (35.3) | 76 (22.6) | 27 (38.5) | 49 (67.1) | 28 (100) | 35 (59.3) | 62 (100) | 7 (53.8) | 208 (68.2) |
| Suction | 1 (0.7) | 1 (0.3) | 9 (12.9) | 5 (6.9) | 0 (0) | 0 (0) | 0 (0) | 1 (7.7) | 15 (4.9) |
| Forceps | 3 (2) | 0 (0) | 9 (12.9) | 3 (4.1) | 0 (0) | 0 (0) | 0 (0) | 0 (0) | 12 (3.9) |
| LSCS | 91 (60.7) | 259 (77.1) | 25 (35.7) | 16 (21.9) | 0 (0) | 24 (40.7) | 0 (0) | 5 (38.5) | 70 (23) |
| Assisted breech | 2 (1.3) | - | - | - | - | - | - | - | - |
| Rotavirus infection<sup>c</sup> |  |  |  |  |  |  |  |  |  |
| Yes | 74 (49.3) | 99 (29.5) | 52 (75.4) | 51 (69.9) | 4 (14.3) | 46 (78) | 5 (8) | 6 (46.2) | 164 (54) |
| No | 76 (50.7) | 237 (70.5) | 17 (24.6) | 22 (30.1) | 24 (85.7) | 13 (22) | 57 (92) | 7 (53.8) | 140 (46) |

<sup>a</sup>Young mothers were defined as <23 years and <27 years of age in the RoVI cohort and NICU-SRMC, Chennai study, respectively.
<sup>b</sup>Missing data for 1 subject in NICU-CMC and 21 subjects in the RoVI cohort.
<sup>c</sup>Unknown for one subject in the RoVI cohort.
Abbreviations: NICU, Neonatal Intensive Care Unit; CMC, Christian Medical College, Vellore; SRMC, Sri Ramachandra Medical College and Research Institute, Chennai; RoVI, Rotavirus Vaccine Immunogenicity study; CHAD, Community Health and Department, Vellore; LCECU, Low Cost Effective Care Unit, Vellore; GVMCH, Government Vellore Medical College and Hospital; UHC, Urban Health Centre; NA, Not Available; LSCS, Lower Segment Caesarean Section.

**TABLE 3.**
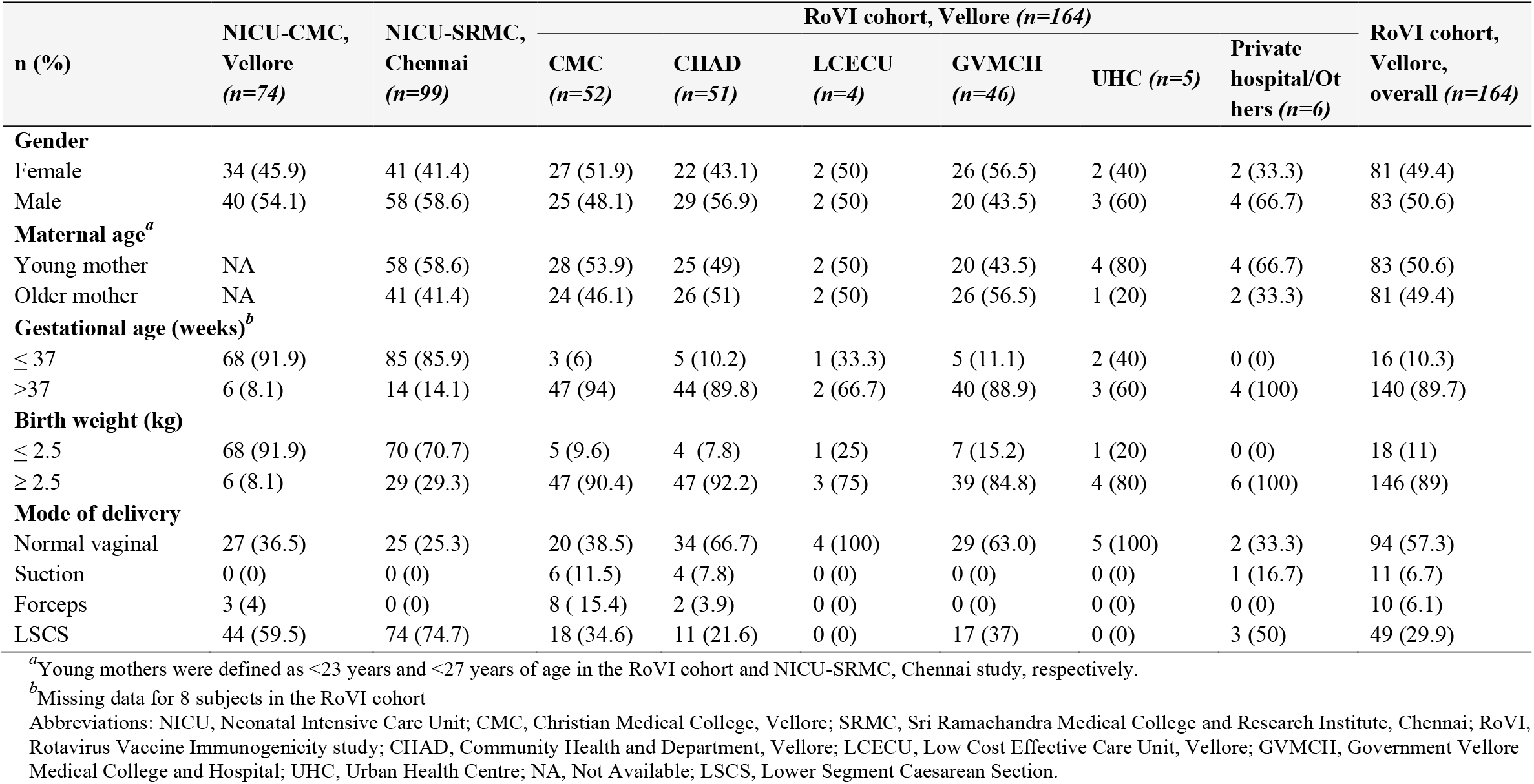
Characteristics of neonates with rotavirus infection

Information regarding the gender of the infant, maternal age, gestational age at birth, birth weight and mode of delivery were collected. Maternal age was classified as young and older mothers using the median cut-off value. Infants born at less than and more than 37 weeks were classified as preterm and term born, respectively. Birth weight was classified as normal if the birth weight was ≥ 2.5 kg. Stool samples were collected approximately within the first four weeks of life from neonates admitted at NICUs of SRMC and CMC. For the RoVI cohort, stool samples were collected at weeks one, four and six of age to determine the persistence of rotavirus infection (Fig. 1).

**FIG 1.**
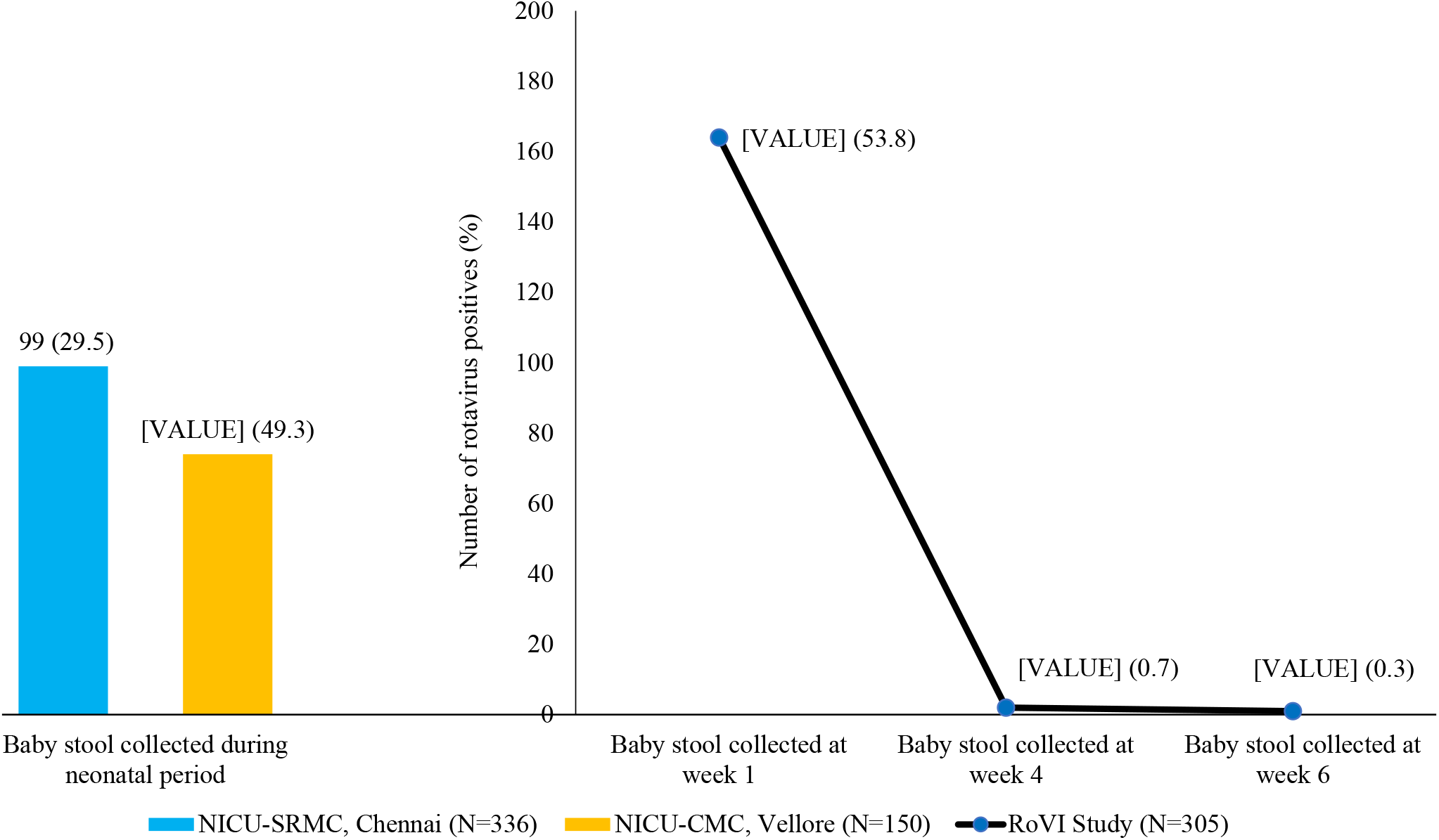
Proportion of infants with rotavirus infection (stool tested using ELISA) from the three study settings NICU: Neonatal Intensive Care Unit; CMC: Christian Medical College, Vellore; SRMC: Sri Ramachandra Medical College and Research Institute, Chennai; RoVI: Rotavirus Vaccine Immunogenicity study

Stool samples collected from the NICUs were tested for the presence of rotavirus antigen using a commercially available ELISA kit (Premier^TM^ Rotaclone®, Meridian Biosciences) as per manufacturer’s instructions. Samples showing an OD value of ≥0.150 were reported as positive for rotavirus antigen. Stool samples collected in the RoVI cohort were tested for rotavirus antigen using an in-house validated ELISA (15). Samples positive in ELISA were further genotyped using a hemi-nested multiplex Polymerase Chain Reaction (PCR).

Nucleic acid was extracted from the positive samples using the QIAamp Viral RNA Mini Kit. Complementary DNA was synthesized using random primers, Pd(N)6 hexamers, and genotypes as previously described using oligonucleotide primers to detect VP7 genotypes G1, G2, G3, G4, G8, G9, G10, and G12 and VP4 genotypes P[4], P[6], P[8], P[9], P[10], and P[11] (16). Alternate primer sets and protocols were used if we failed to genotype the virus by the standard primer sets mentioned (17,18).

Twenty-nine rotavirus positive samples (15 from the RoVI study and 14 from the NICU at CMC) that were PCR confirmed for the G10P[11] strain were selected by random sampling, and subjected to partial sequencing of the VP4 and VP7 encoding genes (876 bp and 881 bp, respectively) and analyzed as described. The first-round amplicons from the genotyping PCR were purified using ExoSAP-IT reagent^®^ (Affymetrix) and sequenced using Big Dye terminator v3.1 cycle sequencing kit (Applied Biosystems) as per the manufacturers’ instructions. The sequences were resolved using an automated sequencer (Genetic Analyzer 3130, Applied Biosystems) and the forward and reverse sequences were assembled using Sequencher V5.4.6 software (Genecodes Corporation) (19). Multiple sequence alignment was performed using MAFFT V7.380 (Multiple Alignment using Fast Fourier Transform) software and edited using the Aliview software (20,21). Phylogenetic analysis was performed using MEGA V7 software and the phylogenetic trees were constructed using the Kimura 2-parameter substitution model with 1000 bootstrap values, visualized and edited using Figtree V1.4.3 software (22,23).

### Accession number(s)

Sequence data for the 29 samples were submitted to the GenBank and assigned accession numbers MN968974-MN969031.

The studies were approved by the Institutional Review Board and Ethics Committee of CMC, Vellore, for the Vellore site, and the Institutional Ethics Committee of SRMC, Chennai for the study at SRMC.

## RESULTS

In total, 791 neonates were included in this study: 150 and 336 neonates admitted for more than 48 hours at the NICU at CMC, Vellore, and NICU-SRMC, respectively, and 305 neonates from the RoVI study (Table 2).

Neonates included from the NICU at CMC, Vellore, were born at a mean gestational age of 32 weeks (SD 3.1; range 24 - 40 weeks), the majority were born preterm (91.3%, 137/150) and 92.7% (139/150) were less than 2.5 kg (range 0.6 - 3.3 kg) (Table 2). Sixty-one percent of the babies (91/150) were born by LSCS with 55.3% (83/150) being male babies. Feed intolerance was the most common diagnosis on admission to the NICU at CMC (49/150, 32.7%). The other associated symptoms were vomiting and vomiting with abdominal distension. One neonate was diagnosed with necrotizing enterocolitis. More than half of the neonates (88/150, 58.7%) did not have any associated gastrointestinal symptoms.

Of the 336 neonates included from NICU-SRMC, Chennai, 55.7% (187/336) were males, born at a mean gestational age of 33 weeks (SD 3.6: range 25 - 40 weeks), with the majority being preterm (287/336, 85.4%) and less than 2.5 kg (246/336, 73.2%) (range 0.6 - 4.4 kg). The mean maternal age was 27 years (SD 4.7, range 18 - 46 years). The majority were born by LSCS (lower segment caesarean section) (259/336, 77.1%).

The 305 neonates from the RoVI cohort were born at a mean gestational age of 38 weeks (SD 1.2; range 34 - 42 weeks), the majority were males (155/305, 50.8%), term-born (257/305, 90.5%) and more than 2.5 kg (269/305, 88.2%) (range 1.8 – 4.2 kg) (Table 2). The mean maternal age was 23 years (SD 3.72; range 17 - 39 years) and the majority were born by normal vaginal delivery (208/305, 68.2%).

Among the 150 neonates recruited from the NICU at CMC, Vellore, 49.3% (74/150) were positive for rotavirus by stool ELISA (Table 3, Figure 1). No significant predisposing factors were identified in neonates from this cohort (Table 4). Of the 74 rotavirus positive neonates, none of them had diarrhea. Feed intolerance (49/74, 66.2%) was the most common gastrointestinal symptom in these neonates. Of the 74 positive samples, 89.2% (66/74) were fully typed. G10P[11] was present in 87.8% (65/74), one sample was genotyped as G12P[11], and eight samples remained untyped.

**TABLE 4.** Comparison of rotavirus infection in the three study settings

| Variable |  | NICU-CMC, Vellore<br>(n=150) |  | p-value | OR<br>(95% CI) | NICU-SRMC, Chennai<br>(n=336) |  | p-value | OR<br>(95% CI) | RoVI cohort, Vellore<br>(n=305) |  | p-value | OR<br>(95% CI) |
| --- | --- | --- | --- | --- | --- | --- | --- | --- | --- | --- | --- | --- | --- |
|  |  | RV<br>Positive<br>n (%) | RV<br>Negative<br>n (%) |  |  | RV<br>Positive<br>n (%) | RV<br>Negative<br>n (%) |  |  | RV<br>Positive<br>n (%) | RV<br>Negative<br>n (%) |  |  |
| **Gender** | Male | 40 (48.2) | 43 (51.8) | 0.76 | 0.9<br>(0.47, 1.71) | 58 (31) | 129 (69) | 0.49 | 1.18<br>(0.74,1.9) | 83 (53.5) | 72 (46.5) | 0.94 | 0.98<br>(0.63, 1.54) |
|  | Female | 34 (50.7) | 33 (49.3) |  |  | 41 (41.4) | 108 (45.6) |  |  | 81 (54) | 69 (46) |  |  |
| **Maternal age<sup>a</sup>** | Young mother | NA | NA | NA | NA | 58 (32.4) | 121 (67.6) | 0.21 | 1.36<br>(0.84, 2.18) | 83 (49.4) | 85 (50.6) | 0.09 | 0.68<br>(0.43, 1.06) |
|  | Older mother | NA | NA |  |  | 41 (26.1) | 116 (73.9) |  |  | 81 (59.1) | 56 (40.9) |  |  |
| **Gestational age (weeks)<sup>b</sup>** | ≤ 37 | 68 (49.6) | 69 (50.4) | 0.98 | 0.98<br>(0.3, 3.2) | 85 (29.6) | 202 (70.4) | 0.88 | 1.05<br>(0.54, 2.06) | 16 (59.3) | 11 (40.7) | 0.64 | 1.22<br>(0.54, 2.72) |
|  | >37 | 6 (50) | 6 (50) |  |  | 14 (28.6) | 35 (71.4) |  |  | 140 (54.5) | 117 (45.5) |  |  |
| **Birth weight (kg)** | ≤ 2.5 | 68 (48.9) | 71 (51.1) | 0.72 | 0.8<br>(0.23, 2.74) | 70 (28.5) | 176 (71.5) | 0.50 | 0.84<br>(0.47, 1.41) | 18 (50) | 18 (50) | 0.63 | 0.84<br>(0.42, 1.69) |
|  | ≥ 2.5 | 6 (54.5) | 5 (45.5) |  |  | 29 (32.2) | 61 (67.8) |  |  | 146 (54.3) | 123 (45.7) |  |  |
| **Mode of delivery** | LSCS | 44 (46.8) | 50 (53.2) | 0.42 | 0.76<br>(0.39, 1.48) | 74 (28.6) | 185 (71.4) | 0.51 | 0.83<br>(0.48, 1.44) | 49 (70) | 21 (30) | **0.002** | 2.43<br>(1.37, 4.31) |
|  | Vaginal | 30 (53.6) | 26 (46.4) |  |  | 25 (32.5) | 52 (67.5) |  |  | 115 (48.9) | 120 (51.1) |  |  |

<sup>a</sup>Young mothers were defined as <23 years and <27 years of age in the RoVI cohort and NICU-SRMC, Chennai study, respectively.
<sup>b</sup>Missing data for 1 subject in NICU-CMC and 21 subjects in the RoVI cohort.
Abbreviations: NICU, Neonatal Intensive Care Unit; CMC, Christian Medical College, Vellore; SRMC, Sri Ramachandra Medical College and Research Institute, Chennai; RoVI, Rotavirus Vaccine Immunogenicity study; RV, Rotavirus; NA, Not Available; LSCS, Lower Segment Caesarean Section.
NICU: Neonatal Intensive Care Unit; CMC: Christian Medical College, Vellore; SRMC: Sri Ramachandra Medical College and Research Institute, Chennai; RoVI: Rotavirus Vaccine Immunogenicity study

Of the 336 neonates from NICU-SRMC, 29.5% (99/336) were positive for rotavirus by stool ELISA (Table 3, Figure 1). No significant factors, to acquire rotavirus infection, were identified among these neonates. (Table 4). Ten neonates (2.9%, 10/99) had associated gastrointestinal symptoms. Of the 99 rotavirus positive samples, 94.9% (94/99) could be fully typed and belonged to the G10P[11] genotype, two stool samples were partially typed-G10P[UT] and three samples were untypable.

Of the 305 neonates from the RoVI study, 54% (164/305) were positive for rotavirus by ELISA during the first week of life (Table 3, Figure 1). By week four and six of age, only two and one infant/s, respectively, were positive for rotavirus. Among the neonates born at different hospitals in Vellore, neonates born at the tertiary care centres (CMC, Vellore, and GVMCH) had the highest rates of infection, 75% (52/70) and 78% (46/59), respectively. With reference to the mode of delivery, neonates in the RoVI cohort showed a significantly higher odds of acquiring a rotavirus infection when the mode of delivery was LSCS when compared to those delivered vaginally [p=0.002, OR=2.4 (1.4-4.3)]. No other factors were found to predispose the neonates for a rotavirus infection (Table 4). None of the neonatal rotavirus infections in the RoVI study was associated with any symptoms. Of the 164 stool samples positive for rotavirus by ELISA, 98.1% (161/164) could be completely typed and belonged to the G10P[11] genotype, one sample was partially typed-G10 P[UT], one had a mixed infection-G1+10, P[1] and P[11] and one sample remained untyped even after using alternate primer sets. At week four of age, two infants had G10P[11] infection, and one infant at week six had a G10P[11] infection. The infants positive at week four of age were infected at week one of age as well. The one infant positive at week six of age was negative for rotavirus at week one and week four of life.

Phylogenetic analysis of the VP4 and VP7 regions were carried out for a total of 29 randomly selected samples that were positive for neonatal rotavirus infection from the RoVI cohort as well as and those previously isolated in Vellore. A high degree of homology was observed between the RoVI cohort strains and the strains isolated previously in Vellore (Figure 2A and 2B).

**FIG 2.**
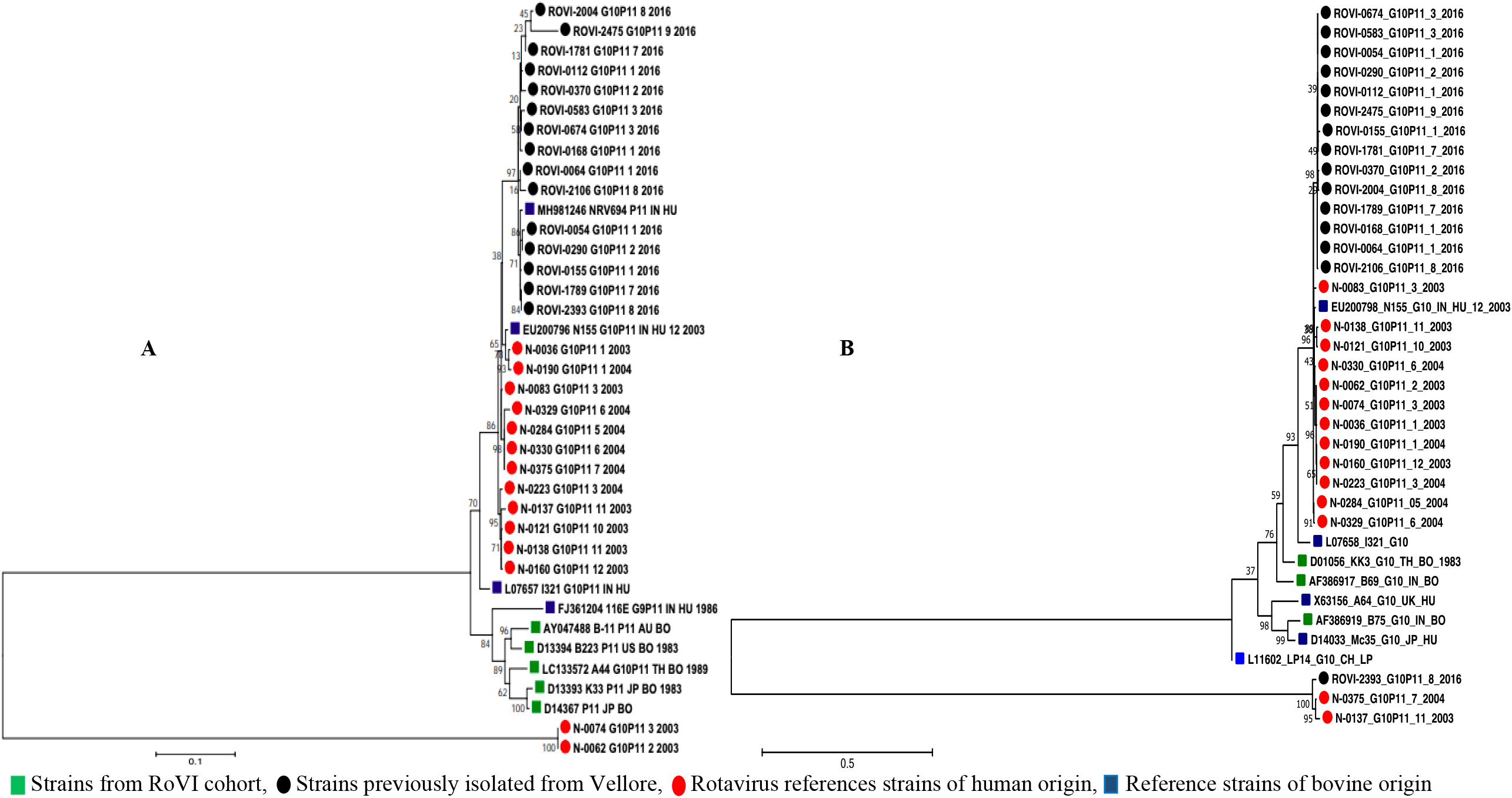
VP4 (Figure 2A) and VP7 (Figure 2B) sequence analysis showing the degree of homology of G10P[11] strains between RoVI cohort and isolates identified previously from Vellore

## DISCUSSION

Linking data from the present study with published data, the persistence of the G10P[11] strains, in neonates born in Vellore, both in the community healthcare setting and the tertiary hospital nursery over the last two decades is evident (4,24). The inclusion of samples collected from a birth cohort allowed us to establish the short duration of this infection, as the infection was cleared in the vast majority of the infants by 4 weeks of age.

This is the first report describing neonatal rotavirus infections in a NICU setting from Chennai. Rotavirus was identified in 29.5% of the neonates admitted, with 2.9% having associated gastrointestinal symptoms. Studies from different parts of the world have reported gastrointestinal symptoms ranging from 13.3% in Korea to 30.4% in Greece, and as high as 43.9% in Vellore among neonates infected with rotavirus (4,25,26). It is interesting to note the difference in rotavirus infection rates between the NICU settings of Vellore and Chennai (49.3% and 29.5%, respectively), showing the difference in the burden of infection between different tertiary care settings.

The neonates enrolled from the CMC and SRMC NICUs did not have any specific risk factors associated with rotavirus infection namely, gender, maternal age, gestational period, birth weight or mode of delivery. This is similar to a previous study conducted in the NICU setting of CMC, Vellore, where the above-mentioned factors showed no association with a neonate acquiring either a symptomatic or asymptomatic rotavirus infection (4). Also, a similar study from Bangladesh did not demonstrate any significant factors predisposing neonates to acquire rotavirus infection (27). However, in contrast, a study from Madrid showed that prematurity and lower birth weight predisposed neonates to rotavirus infection in a nursery setting, though our study showed no such association (10). The association between neonatal rotavirus infection and the mode delivery with reference to vaginal delivery or caesarean section has not been clearly established. In a study from Australia normal vaginal delivery with the early establishment of breastfeeding reduced the risk of acute gastroenteritis related hospitalization in young children, though this was not rotavirus specific (28). In the RoVI cohort, neonates born by LSCS had a higher risk of neonatal rotavirus infection. The majority of the neonates born by LSCS are usually kept under observation before being roomed-in with the mother. This suggests that even a short duration of stay by the new-born in the nursery could increase the risk of acquiring a rotavirus infection. A previous study from Vellore has documented the presence of G10P[11] rotavirus in the environmental swabs taken from the nurseries (4,29).

Prior studies from Delhi, India, reported a 46% reduction in subsequent rotavirus infections in infants who had a neonatal rotavirus infection (5,30). The data from Delhi lead to the subsequent development and licensure of the Rotavac^®^ vaccine based on the neonatal strain G9P[11]. Clinical trials using a neonatal dosing regimen involving Rotavac^®^ and RV3-BB, another vaccine developed from a neonatal strain G3P[6], are currently underway (31,32). In contrast, previous studies in Vellore showed that neonatal rotavirus infection did not provide any subsequent protection from rotavirus diarrhea of any severity (6). Despite this, it will be important to analyse any potentially enhancing or inhibitory effect of the G10P[11] neonatal strain on vaccine take and vaccine-induced immunity.

The sustained persistence of the G10P[11] neonatal strains in Vellore contrasts with the findings from studies from Delhi in northern India. Previous reports from Delhi showed the predominance of G9P[11] strain between 1986 to 1992, which was replaced by a second novel strain, G9P[6] in 1993 (33). Between 2005-2006, G12P[6] strain was described as the dominant strain among neonates in nurseries in Delhi (34). Reasons for the persistence of the G10P[11] rotavirus strain among neonates in Vellore are not fully understood, but available data suggest host-virus co-evolution; the role of the HMOs in enhancing infectivity, and the age-restricted tropism exhibited by G10P[11] through binding to H-antigen glycan precursors (13,35).

For rotavirus infection to establish in the gut, the VP8 domain of the VP4 spike protein binds to specific glycans. The association of G10P[11] with infection in neonates has been explained by the unique specificity of the spike domain to host glycans that are restricted to the neonatal period. Recent crystallographic studies have demonstrated that the bovine component of the strain P[11] VP8 domain binds specifically to type II glycans found within the bovine gut (35,36). However, the strain binds to both type I and type II precursor glycans found abundantly in the early neonatal gut as well as the human breastmilk. The VP8 region is the least conserved region among all rotavirus structural proteins. It may gradually assimilate sequence changes, leading to the zoonotic crossover to humans in the neonatal period. The presence of these glycans predominantly in the neonatal gut is known and the assessment of the effect of HMOs and receptor tropism of the neonatal strains should help elucidate the factors related to the persistence of the G10P[11] strains.

The main strength of this study was that we were able to compare neonatal rotavirus infection in neonatal intensive care settings of two tertiary care hospitals and a community-based birth cohort. The addition of the community cohort is an important highlight in this study, showing a high burden of asymptomatic neonatal rotavirus infection in the community with the persistence of the G10P[11] strain in the NICU settings of both hospitals. A limitation of the study was the non-availability of data on the time interval between the delivery and initiation of breastfeeding in the new-born infants. Further, no data was available on the duration of the stay at the hospital for individual mothers though the duration of hospital stay for a mother who underwent a caesarean section is longer than for vaginal delivery.

In conclusion, this study highlights the widespread nature and high transmissibility of the rotavirus genotype G10P[11] and its continued persistence among neonates born in diverse health care settings across Vellore and in Chennai. We show the persistence of the virus over the last two decades in the nursery at Vellore. We note the virus was not restricted to the large hospital nurseries but widely prevalent in the community hospitals in Vellore. These data highlight the importance of surveillance in neonates for rotavirus infections, even without gastrointestinal symptoms, as these may impact subsequent rotavirus vaccine response.

## Data Availability

All data can be made available on written request

## ACKNOWLEDGEMENTS

The work at Christian Medical College, Vellore was funded by DBT (Department of Biotechnology (grant number BT/IN/DBT-MRC/DFID/23/GK/2015-16 dated 11.12.2015 to G.K.) India. The work was also supported by the National Institutes of Health (R01AI105101 to M.K.E., S.R., L.B. and G.K. The work at SRMC (Sri Ramachandra Medical College), Chennai was funded by ICMR-TSS (Indian Council of Medical Research -Talent Search Scheme).

The funding agency had no role in study design, interpretation, or the decision to submit the work for publication.

We have no conflicts of interest to report.

We thank the community, mothers and their infants of Chinnallapuram. The authors thank Mr Annai Gunasekaran who supervised the field activities for the study at Vellore. We thank the staff of Sri Ramachandra Medical College and Research Institute for coordinating the study.

